# Non synonymous substitutions in HIV-1 GAG are frequent in epitopes outside the major hydrophobic region and associated with subtype differences

**DOI:** 10.1101/2020.04.09.20057703

**Authors:** Babatunde A. Olusola, David O. Olaleye, Georgina N. Odaibo

## Abstract

In 2018, an estimated 38 million people lived with HIV-1 infection resulting in 770,000 deaths. More than 50% of this infection and its associated deaths occurred in Sub-Saharan Africa. There is the need to develop an HIV-1 vaccine if the epidemic would be effectively controlled. Cytotoxic T Lymphocytes (CTL) epitopes within the Major Hydrophobic Region (MHR) have been shown to be highly immunogenic and immuno-dominant. These conserved epitopes have recently been the focus of vaccines studies. Despite the West African epicenter having one of the highest numbers of diverse circulating HIV-1 strains, very few longitudinal studies have checked the frequencies of CTL immune escape variants on epitopes within and without the MHR for HIV-1 strains circulating in the region.

In this study we describe non-synonymous substitutions within and without the MHR of HIV-1 GAG genes isolated from 10 infected Nigerians at the early stages of HIV-1 infection. Furthermore, we analyzed these substitutions longitudinally in five infected individuals from the early stages of infection up until when antibodies become detectable. We identified 3 non synonymous substitutions within the MHR of HIV-1 GAG genes isolated from the early HIV infected individuals. Fourteen and nineteen mutations outside the MHR were observed before and after detection of antibodies respectively while 4 mutations were found within the MHR. These substitutions include previously mapped CTL epitope immune escape mutants. CTL immune pressure likely leave different footprints on HIV-1 GAG epitopes within and outside the MHR. This information is crucial for future vaccine designs for use in the West African region.

## Introduction

In 2018, an estimated 38 million people lived with HIV-1 infection resulting in 770,000 deaths. The rate of new infections also increased in the Sub-Sahara Africa region. More than 50% of HIV infection and its associated deaths also occurred in this region[1]. Combination antiretroviral therapies (cART) have been effective in suppressing viremia to undetectable levels, increasing survival, improving quality of life and decreasing infectiousness in infected individuals[2,3]. However, just about half of infected individuals receive treatment. Sub-Sahara Africa despite being the highest hit region with the virus, has the lowest access to treatment[1]. Access to treatment is unlikely to increase to optimal levels because of economic, social and pharmacologic challenges associated with cART use[2,4]. Moreover, cART is not curative as persons on treatment has to use drugs to control emergence of latent reservoir HIV (immune escape as well as drug resistant strains) for the rest of their lives[2,5]. Therefore, to control the HIV epidemic especially in African countries, there is the need to develop a safe, cost-effective, durable and accessible HIV-1 vaccine[4,6–8].

Induction of broadly neutralizing antibodies is the main stay of all protective anti-viral vaccines. However, this has been very difficult to generate for HIV-1 because of the plasticity of the virus genes due to the action of reverse transcriptase enzyme[2,4,7]. Also, broadly neutralizing antibodies develop very late in infection after establishment of the latent reservoir strains[9,10]. Recent vaccine efforts have been directed towards the stimulation of cellular immunity. Therefore, consideration of HIV-1 vaccine candidates that stimulate cellular immunity has been the focus of many recent vaccine studies. Cytotoxic T Lymphocytes (CTL) commonly referred to as CD8+ T cells have been extensively showed to control HIV-1 replication especially during the early stages of infection[4,11,12]. Previous studies have associated expansion of CTL with the control of acute infections[13–17].

Although non-synonymous substitutions are more predominant in the ENV gene of HIV-1, studies have shown that these substitutions also occur in CTL epitopes of the HIV-1 GAG gene especially in the P17 and P24 regions[18–21]. Initially, CTL control of acute HIV infections were associated with HLA protection[22–25], recent studies have however showed that magnitude and breadth of CTL responses as well as CTL epitopes presented by similar HLA alleles during the early stages of infection may better predict disease outcomes[26–32]. CTL epitopes within the major hydrophobic region (MHR) have been shown to be promiscuously presented by similar HLA alleles[33]. The MHR, found within the capsid gene, is a conserved motif among retroviruses[22,34]. This region is important in particle assembly and viral infectivity[35]. Furthermore, CTL epitopes within the MHR have been shown to be highly immunogenic and immuno-dominant [19–21,36]. It is also hypothesized that escape mutations within the MHR are likely to be deleterious to HIV as they seem to be associated with fitness cost[37–39]. These conserved epitopes (Gag 240–249, TSTLQEQIGT; Gag 162–172, KAFSPEVIPMF and Gag 203–212, ETINEEAAEW) have recently been the focus of universal therapeutic vaccines[2,6,7,19,40].

However, there is limited information on immune escape strains within and without the MHR for HIV-1 strains circulating in the West African epicenter. Immune Escape due to CTL epitopes outside the MHR may be a crucial factor to consider in the design of therapeutic HIV vaccines[41] as they may provide opportunities for compensatory mutations on replicative fitness[42]. It is also important for HIV-1 vaccines when developed, to be effective against strains circulating in African countries[4]. Despite the West African epicenter having one of the highest numbers of diverse circulating HIV-1 strains, very few longitudinal studies have checked the frequencies of CTL immune escape variants on epitopes within and without the MHR for HIV-1 strains circulating in the region. In this study we describe non-synonymous substitutions within and without the MHR of HIV-1 GAG genes isolated from 10 infected Nigerians at the early stages of HIV-1 infection. These individuals were part of a previously described study[43] (and study 3). Furthermore, using phylogenetic tools as well as programs and databases in the Los Alamos National Laboratory HIV Sequence Database (https://www.hiv.lanl.gov), we analyze these substitutions longitudinally in five infected individuals from the early stages of infection up until when antibodies become detectable.

## Methods

### Study sites and patient population

Twenty-three individuals at the early stages of HIV-1 infection identified and described previously (study3) were recruited for this study (see Table 1). Ten persons out of these were studied for early HIV-1 infection. Another 5 individuals identified in 2017 were followed up until antibodies were detected. Profile of the follow up schedule is shown in Table 2. HIV-1 antibodies were detected after two years, although the exact time when HIV-1 antibodies became detectable is unknown. There was about 12 month’s interval between the penultimate visit and the one in which antibodies were detected. At every visit, these individuals were screened for HIV-1 infection using earlier described protocol[43,44].

**Table 1:**
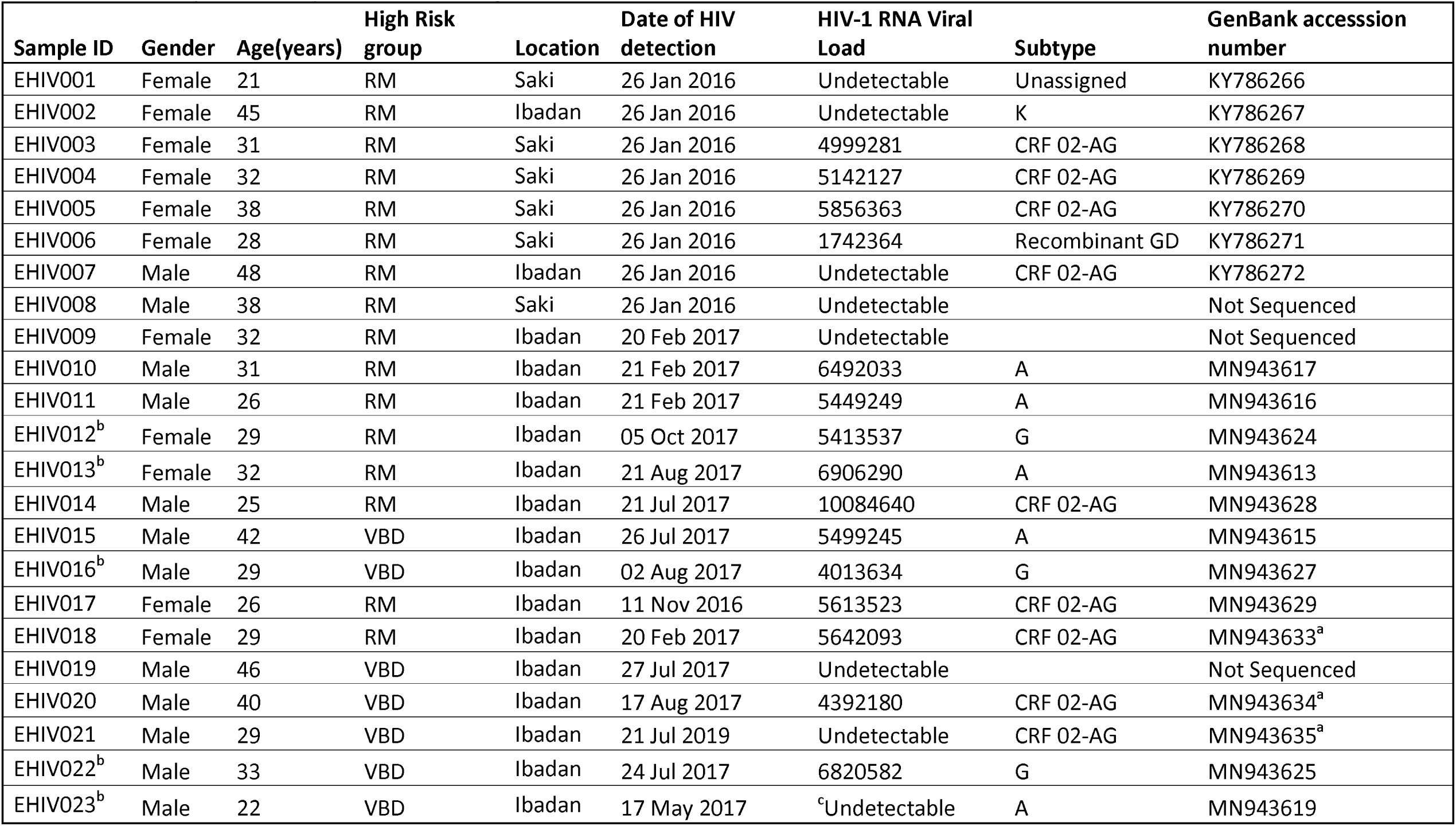
Summary on early HIV-1 infected Nigerians. Individuals comprised of persons referred for malaria parasite testing (RM) and Voluntary Blood Donors (VBD). All blood donors were from Ibadan. ^a^These samples were genotyped using the VIF gene as we did not obtain GAG gene for these individuals. ^b^These individuals were followed up until HIV-1 antibodies were detectable in their sera. ^c^EHIV 023 HIV-1 RNA viral load became detectable after HIV-1 antibodies became detectable.

| Sample ID | Gender | Age(years) | High Risk group | Location | Date of HIV detection | HIV-1 RNA Viral Load | Subtype | GenBank accession number |
| --- | --- | --- | --- | --- | --- | --- | --- | --- |
| EHIV001 | Female | 21 | RM | Saki | 26 Jan 2016 | Undetectable | Unassigned | KY786266 |
| EHIV002 | Female | 45 | RM | Ibadan | 26 Jan 2016 | Undetectable | K | KY786267 |
| EHIV003 | Female | 31 | RM | Saki | 26 Jan 2016 | 4999281 | CRF 02-AG | KY786268 |
| EHIV004 | Female | 32 | RM | Saki | 26 Jan 2016 | 5142127 | CRF 02-AG | KY786269 |
| EHIV005 | Female | 38 | RM | Saki | 26 Jan 2016 | 5856363 | CRF 02-AG | KY786270 |
| EHIV006 | Female | 28 | RM | Saki | 26 Jan 2016 | 1742364 | Recombinant GD | KY786271 |
| EHIV007 | Male | 48 | RM | Ibadan | 26 Jan 2016 | Undetectable | CRF 02-AG | KY786272 |
| EHIV008 | Male | 38 | RM | Saki | 26 Jan 2016 | Undetectable |  | Not Sequenced |
| EHIV009 | Female | 32 | RM | Ibadan | 20 Feb 2017 | Undetectable |  | Not Sequenced |
| EHIV010 | Male | 31 | RM | Ibadan | 21 Feb 2017 | 6492033 | A | MN943617 |
| EHIV011 | Male | 26 | RM | Ibadan | 21 Feb 2017 | 5449249 | A | MN943616 |
| EHIV012<sup>b</sup> | Female | 29 | RM | Ibadan | 05 Oct 2017 | 5413537 | G | MN943624 |
| EHIV013<sup>b</sup> | Female | 32 | RM | Ibadan | 21 Aug 2017 | 6906290 | A | MN943613 |
| EHIV014 | Male | 25 | RM | Ibadan | 21 Jul 2017 | 10084640 | CRF 02-AG | MN943628 |
| EHIV015 | Male | 42 | VBD | Ibadan | 26 Jul 2017 | 5499245 | A | MN943615 |
| EHIV016<sup>b</sup> | Male | 29 | VBD | Ibadan | 02 Aug 2017 | 4013634 | G | MN943627 |
| EHIV017 | Female | 26 | RM | Ibadan | 11 Nov 2016 | 5613523 | CRF 02-AG | MN943629 |
| EHIV018 | Female | 29 | RM | Ibadan | 20 Feb 2017 | 5642093 | CRF 02-AG | MN943633<sup>a</sup> |
| EHIV019 | Male | 46 | VBD | Ibadan | 27 Jul 2017 | Undetectable |  | Not Sequenced |
| EHIV020 | Male | 40 | VBD | Ibadan | 17 Aug 2017 | 4392180 | CRF 02-AG | MN943634<sup>a</sup> |
| EHIV021 | Male | 29 | VBD | Ibadan | 21 Jul 2019 | Undetectable | CRF 02-AG | MN943635<sup>a</sup> |
| EHIV022<sup>b</sup> | Male | 33 | VBD | Ibadan | 24 Jul 2017 | 6820582 | G | MN943625 |
| EHIV023<sup>b</sup> | Male | 22 | VBD | Ibadan | 17 May 2017 | <sup>c</sup>Undetectable | A | MN943619 |

**Table 2:**
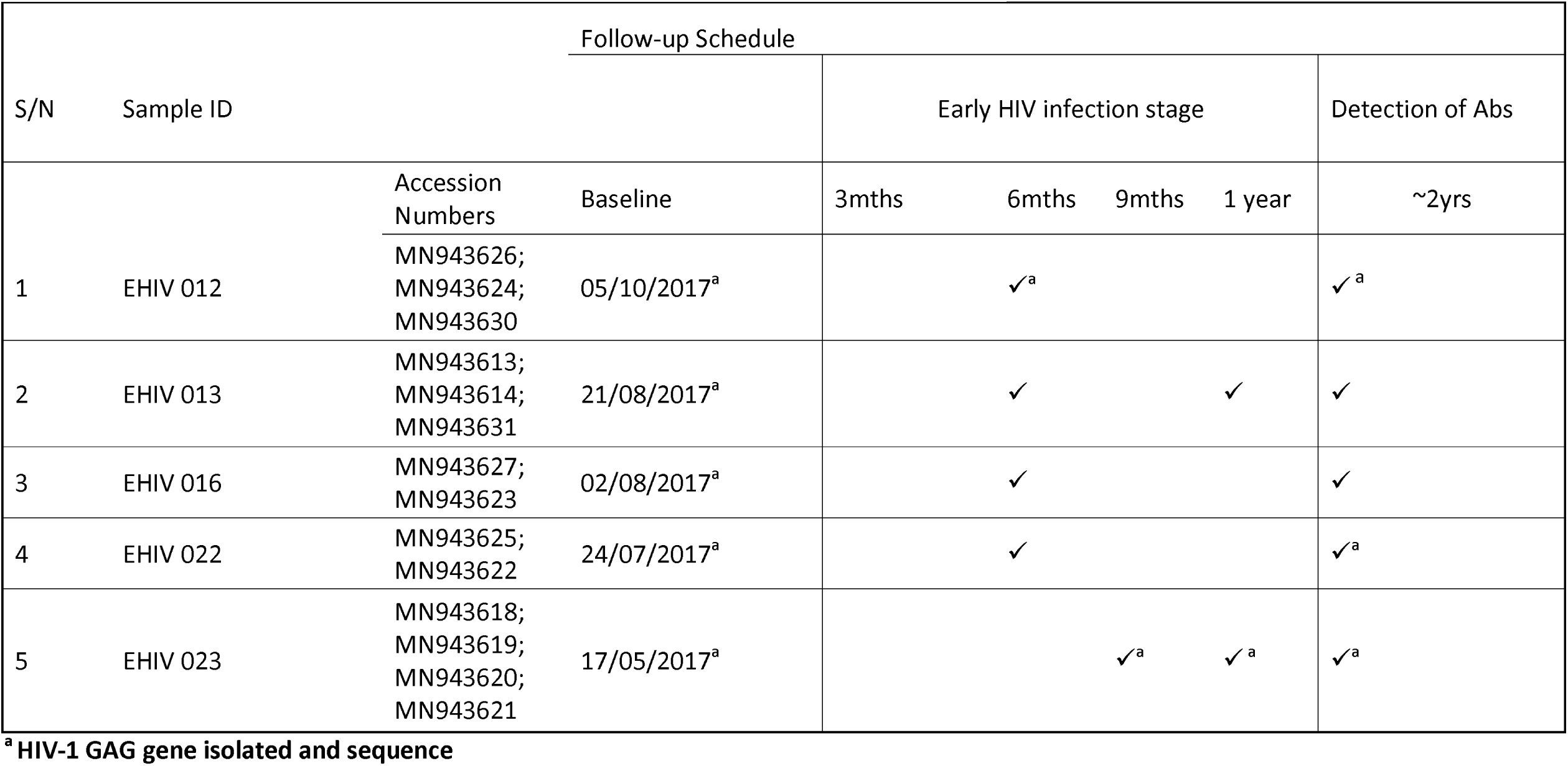
Analysis of samples collected during follow up. Blood samples collected from the five individuals were analyzed for HIV-1 GAG gene. All samples were analyzed at baseline, after HIV-1 antibodies detection and at every time point indicated with superscript a. Each analyzed gene was sequenced and successfully submitted to GenBank with the accession numbers provided. Each accession number corresponds to a GAG gene sequenced at a particular time point. Two samples were sequenced at two and three time points each while a sample was sequenced at four time points. The time at which HIV-1 antibodies were detected does not correspond to seroconversion date as most of these individuals are likely to have seroconverted before this period.

### Recruitment of participants, sample collection and processing

Participants were recruited for this study after obtaining informed consent. Experiments were conducted with the understanding and the consent of participants. Socio demographic data were also collected from these persons. Feedback on results were provided within a week of sample collection. Individuals were counselled and encouraged to continue presentation for testing. These persons were not placed on ART since they were still at the early stages of infection. In Nigeria, only persons with detectable antibodies are placed on treatment. Five milliliters of whole blood were collected in EDTA bottles from participants at every visit. Plasma was separated from the samples immediately after collection, stored at -20°C and transported in cold chain to a central laboratory for analysis. The samples were then stored at -80°C until analyzed. Blood samples were analyzed for HIV antigen/antibody, serum creatinine, HIV-1 RNA viral load (at baseline) and HIV-1 GAG DNA.

### Identification of early HIV-1 infection and detection of antibodies

The updated CDC algorithm of laboratory testing for the diagnosis of early and chronic HIV-1 infection was used for this study as previously described[43]. Briefly, samples were identified as early HIV infection if HIV-1 DNA was detected after being positive with a 4th generation HIV ELISA (detection of both antigen and antibody) and negative with 3^rd^ generation HIV ELISA (detection of HIV antibody only) while those from which HIV-1 DNA scored positive with both 4^th^ and 3^rd^ generation ELISA, were classified as chronic HIV infection. Genscreen™ ULTRA HIV-Ag-Ab ELISA kit (Biorad, Hercules, California) was used as the 4^th^ generation ELISA kit while AiD™ anti-HIV 1+2 ELISA kit (Wantai, Beijing, China) was used as the 3^rd^ generation ELISA kit. All assays were performed under strict biosafety conditions and according to the manufacturer’s recommendation. Furthermore, detection of HIV antibodies were further confirmed using Alere™ Determine™ HIV-1/2 Rapid Antibody Test kit (Alere, Chiba, Japan).

### Clinical chemistry assay for serum creatinine

Plasma samples were analyzed on a Roche cobas® C11 blood chemistry analyzer (Roche Diagnostics, Indianapolis). Each sample was analyzed to determine the level of serum creatinine according to the manufacturer’s instruction. Normal reference ranges for plasma creatinine is 62-133 µmol/L.

### HIV-1 RNA Viral load testing

Serum samples collected at baseline were tested for Plasma HIV-1 viral load (copies/ml) using the COBAS^®^ Ampliprep/COBAS TaqMan96^®^HIV-1 Test, v2:0 (Roche Molecular Diagnostics, Branchburg, NJ, USA) according to manufacturer’s instruction or by an in house real-time PCR protocol.

### PCR amplification and sequencing

Total DNA was extracted from whole blood samples collected at each visit using guanidium thiocyanate in house protocol. A fragment of the gag-pol region (900 base pairs) of the virus was amplified using previously published primers and cycling conditions (Gall 2012)[45] with slight modifications. Briefly, PCR was performed using a SuperScript III One-step RT-PCR system with platinum TaqDNA High fidelity polymerase (Jena Bioscience). Each 25µl reaction mixture contained 12.5µl reaction mix (2x), 4.5µl RNase-free water, 1µl each of each primer (20pmol/µl), 1µl Superscript III RT/Platinum Taq High Fidelity mix, and 5µl of template DNA. Pan-HIV-1_1R (CCT CCA ATT CCY CCT ATC ATT TT) and Pan-HIV-1_2F (GGG AAG TGA YAT AGC WGG AAC) were used. Cycling conditions were 94°C for 5min; 35 cycles of 94°C for 15s, 58°C for 30s, and 68°C for 1min 30s; and finally, 68°C for 10mins. Positive HIV samples that was undetectable using the above stated primers were retested using another set of GAG primers for nested PCR as described previously[46]. Positive PCR reactions were shipped on ice to Macrogen, South Korea for Big Dye sequencing using the same amplification primers (Pan-HIV-1_1R and Pan-HIV-1_2F; or G60 and G25).

### Detection of HIV-1 subtypes and phylogenetic analysis

The sequences were cleaned and edited using Chromas and Bioedit softwares. Subtyping was performed using a combination of four subtyping tools: The Rega HIV-1 Subtyping Tool, version 3.0 (http://dbpartners.stanford.edu/RegaSubtyping/), Comet, version 2.2 (http://comet.retrovirology.lu), National Center for Biotechnology Information, Bethesda, MD (http://www.ncbi.nlm.nih.gov/Blast.cgi) and jpHMMM: Improving the reliability of recombination prediction in HIV-1 (http://jphmm.gobics.de/submission_hiv). The first three tools were used simultaneously while jpHMMM was used to resolve discordant subtypes. Phylogenetic analyses were performed using MEGA software version 10. Alignment of sequences were performed using MAFFTS online software. Genetic distances were inferred using the Tamura-Nei model and a phylogenetic tree was generated using the maximum likelihood method. The robustness of the tree was evaluated with 1000 bootstrap replicates. All consensus nucleotide sequences obtained in this study were submitted to GenBank Database and assigned accession numbers MN943617-635.

### Non-synonymous substitutions in Cytotoxic T Lymphocytes (CTL) epitopes within the Major Hydrophobic Region (MHR) of HIV-1 GAG gene isolated from 10 early infected individuals

Reference GAG sequences for subtypes G, A and CRF02-AG were downloaded from the Los Alamos National Laboratory HIV Sequence Database (https://www.hiv.lanl.gov/content/sequence/NEWALIGN/align.html). Deduced amino acid (aa) sequences were translated for both reference and sample sequences with the standard genetic code using Bioedit software. CTL epitope corresponding to the three highly conserved sites (HCS) of the MHR namely KAFSPEVIPMFSALSEGATPQD, DTINEEAAEWDR and TSTLQEQIR[2,6,19] were used for comparison and identification of amino acid substitution. HIV-1 GAG sequences identified as subtypes A, G and CRF02_AG in this study were aligned with Reference A(GenBank accession numbers DQ676872; AB253421 and AB253429), G(GenBank accession numbers AF084936; AF061641; U88826 and AY612637) and CRF02_AG(GenBank accession numbers L39106 and DQ168578) sequences respectively.

### Non-synonymous substitutions in Cytotoxic T Lymphocytes (CTL) epitopes outside the Major Hydrophobic Region (MHR) of HIV-1 GAG gene

The Virus Epidemiology Signature Patterns Analysis (http://www.hiv.lanl.gov/content/sequence/VESPA/vespa.html) program was used to identify variations in other sites of the HIV-1 GAG sequence outside the HCS corresponding to CTL epitope regions. Deduced amino acid (aa) sequences were translated for both reference and sample sequences with the standard genetic code using Bioedit. HIV-1 GAG sequences identified as subtypes A, G and CRF02_AG in this study were aligned with Reference A (GenBank accession numbers DQ676872; AB253421 and AB253429), G (GenBank accession numbers AF084936; AF061641; U88826 and AY612637) and CRF02_AG (GenBank accession numbers L39106 and DQ168578) sequences respectively. Only amino acid replacements in which there was 100% non-synonymous substitution between reference sequences and sample sequences were considered. Percentage substation rates were calculated by finding the ratio of number of substitutions to the total possible substitution sites. These mutations were compared with the Los Alamos National Laboratory HIV Immunology Database for CTL/CD8+ Epitope Variants and Escape Mutations (https://www.hiv.lanl.gov/content/immunology/variants/ctl_variant.html).

### Ethical approval

This research was conducted in accordance with the declaration of Helsinki. Experiments were conducted with the understanding and the consent of each participant. Ethical approvals for this research were obtained from the University of Ibadan/ University College Hospital (UI/UCH) Research and Ethics Committee (UI/EC/15/0076) and the Oyo State Ministry of Health Committee on Human Research (AD13/479/951). All results were delinked from patient identifiers and anonymized.

### Eligibility/ Exclusion criteria

Only individuals between 18 and 65 years of age were included in the study. Individuals who already knew their HIV status were excluded from the study.

### Data management and statistical analysis

Statistical analyses were performed using SPSS version 20. Data are expressed as means ± standard deviations. Statistical significance was estimated using the Kruskal-Wallis test, with SPSS package version 12.0. Statistical significance was defined as P values \0.05.

## Results

### Participants’ characteristics

As described previously, twenty three individuals were identified to be at the early stages of HIV-1 infection. Figure 1 shows the phylogeny of HIV-1 subtypes. Out of the 10 early infected persons studied, 5 were infected with Subtype A, 3 with subtype G and the rest CRF02-AG. Five individuals at the early stages of HIV-1 infection were followed up until detection of antibodies. These individuals were identified to be at the early stages of infection at different periods in 2017. One individual in October, July and May each and two in August. Three of the individuals were males and were voluntary blood donors. The remaining two females were identified when referred for malaria antigen test. Three of the individuals were infected with HIV-1 subtype G while the other two were infected with subtype A (Table 1).

**Figure 1:**
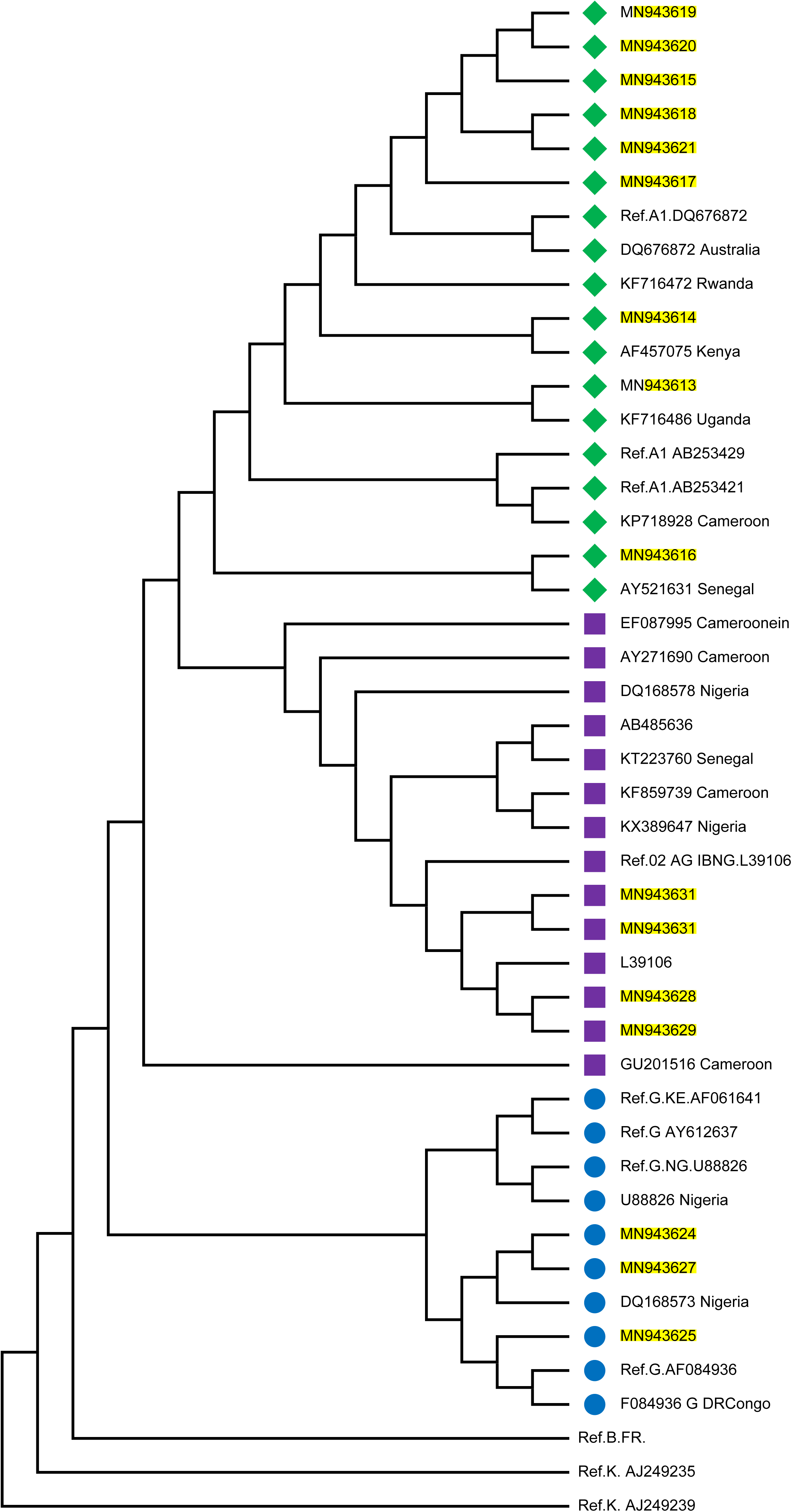
Phylogenetic tree of the P17/P24 regions of the GAG gene of HIV-1. Reference subtypes are indicated with Ref. before their accession numbers. Other sequences are indicated with their accession numbers and country of isolation. Subtypes obtained from samples in this study are indicated with their accession numbers only. Subtypes A, G and CRF02_AG were identified with green diamond, blue circle and pink square symbols respectively. Multiple sequences alignment and phylogenetic tree were constructed using MAFFTS and Maximum Parsimony algorithm in MEGA 6 software. Statistical significance of the tree topology was tested by 1000 bootstrap replication.

Table 2 shows the analysis of the samples collected from these individuals from baseline until after detection of antibodies, two years after. In four individuals, blood samples were collected at three time points, twice during the early stages of infection and once after detection of antibodies. Samples were collected four times in the fifth individual, thrice during the early stages of infection and once after detection of antibodies. HIV-1 GAG sequences of these infected individuals were determined at every time point of blood collection.

### Phylogenetic analysis

Figure 1 shows the estimated phylogeny of HIV-1 subtypes with respect to reference sequences available in the HIV Los Alamos National HIV Sequence database. As shown in the Figure, Subtypes A, G and CRF02_AG were identified with green, blue and pink symbols respectively. HIV-1 subtypes A identified in this study were closely related to Ref A1 DQ676872 as well as subtypes AF457075, KF716486 and AY521631 from Kenya, Uganda and Senegal respectively. Those identified as subtypes G and CRF02-AG were closely related to the Nigerian subtypes DQ168573 and Ref.02 AG IBNG. L39106 respectively.

### There is high rate of substitutions in HCS of CTL epitope regions of Subtype A HIV-1 GAG gene during early infection

In this study we compared intra and inter variations among 10 HIV-1 GAG sequences isolated from persons at the early stages of infection. These sequences were grouped by subtypes and analyzed alongside reference sequences. As shown in Table 3, variations occurred mostly in HIV-1 Subtype A at the CTL epitope region of 243-251aa. In fact, the conserved epitope of TSTLQEQIR was not found in both the reference subtypes and those from early infected individuals. HIV-1 Subtype A also had the highest variations (50%) for HCS corresponding to CTL epitope region (203-214aa). HCS for CTL epitope region 162-183aa was the most conserved among the subtypes, yet a substitution rate of 2.7% was observed for HIV-1 subtype A. CTL escape region (162-183) KAFSPEVIPMFSALSEGATPQD had lowest frequency of mutations. However, two samples (MN943615; MN943616) had two mutations-K162R and A163G. MN943615 had these two mutations while MN943616 had A163G only. These two mutations have been previously associated with immune escape (non-susceptible forms) on the KAFSPEVIPMFSALSEGATPQD epitope[47]. D203E substitution found in a sample (MN943625) in subtype G seems to be a reversion[41].

**Table 3:**
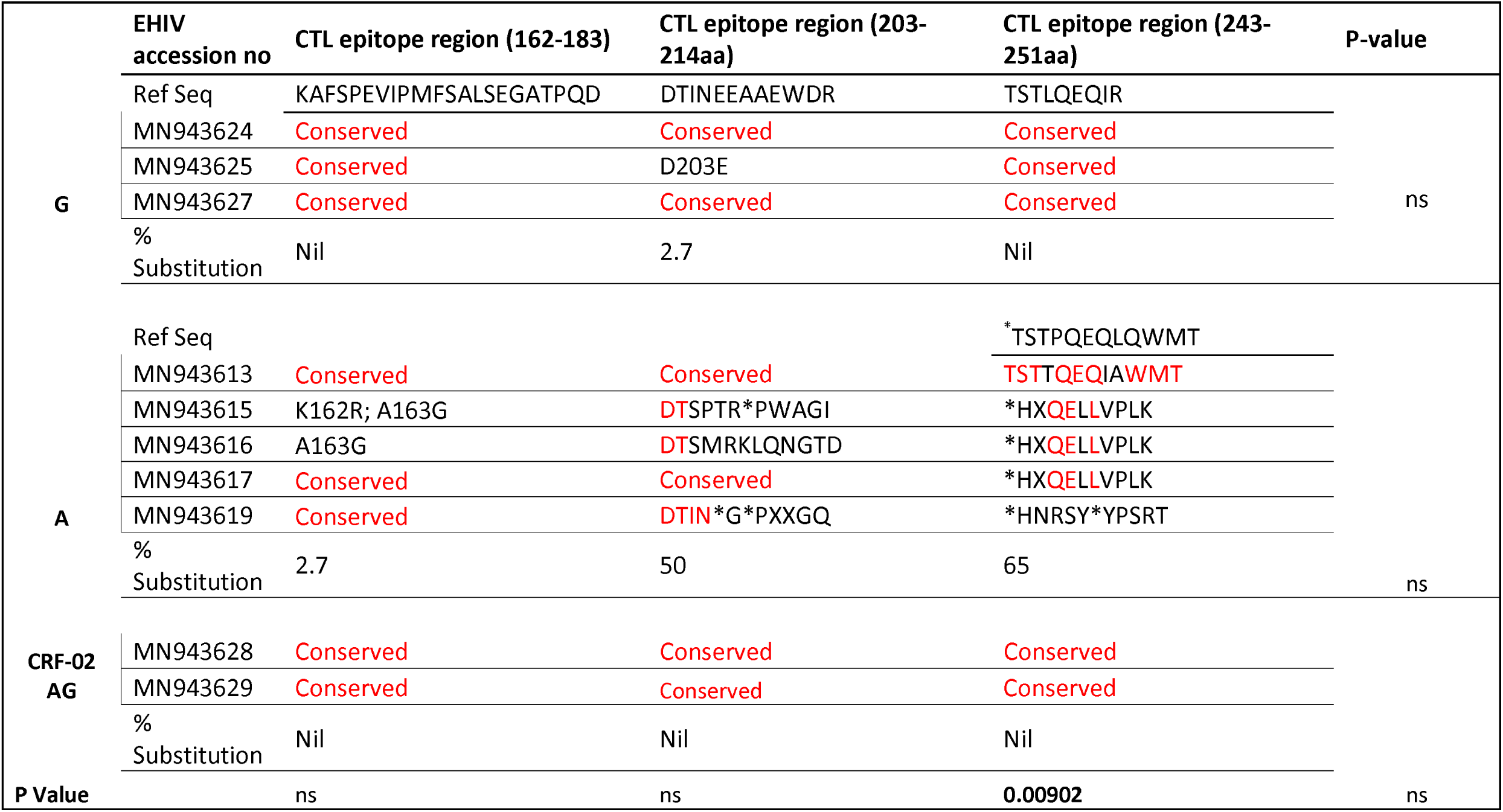
Substitutions associated with highly conserved sites in HIV-1 GAG gene during early HIV infection. Amino acid lengths of CTL epitope regions are indicated in parenthesis. Reference HIV-1 GAG sequences for subtypes G, A and CRF02-AG were downloaded from Los Alamos national HIV sequence database (www.hiv.lanl.gov/content/sequence/NEWALIGN/align.html) and were used for comparisons with similar subtypes obtained from persons at the early stages of HIV infection. Sequences that are not different from reference sequences (presence of the consensus amino acid) are indicated with the word Conserved. If an amino acid position is variable to the reference sequence, the letter and the position is indicated in the normal convention, e.g MN943625 sequence is variable at position 203 and it is indicated as D203E which means at position 203, D is substituted with E. When more than two amino acid positions are variable, the entire sequence is written against the accession number of the subtype. ^*^Reference sequence for subtype A CTL epitope region 243-251 is different from those of other subtypes and it is indicated. %Substitution was calculated by deducing the percentage of the ratio of variable positions against total amino acid positions for each sequence. Statistical tests of significance within and across CTL epitope regions for the subtypes studied were calculated using Kruskal-Wallis test, with P set at 0.05. ns=not significant. Significant values are indicated in bold values.

### Sequences isolated after detection of antibodies are associated with a higher rate of substitutions in HCS of CTL epitope regions

Substitutions in the HCS of subtype B HIV-1 GAG gene have been shown to come with a fitness cost. These substitutions have shown to be deleterious to HIV-1 strains eventual survival and transmission. However, very limited information exist on substitutions associated with immune epitopes during early non-subtype B HIV-1 infection. We followed five individuals from early stage of infection till detection of antibodies. Three of these individuals were infected with subtype G while the remaining two subtype A (see Table 4). There were no substitutions in CTL epitope regions for EHIV016 while EHIV022 had single aa substitutions before and after antibodies detection.

**Table 4:**
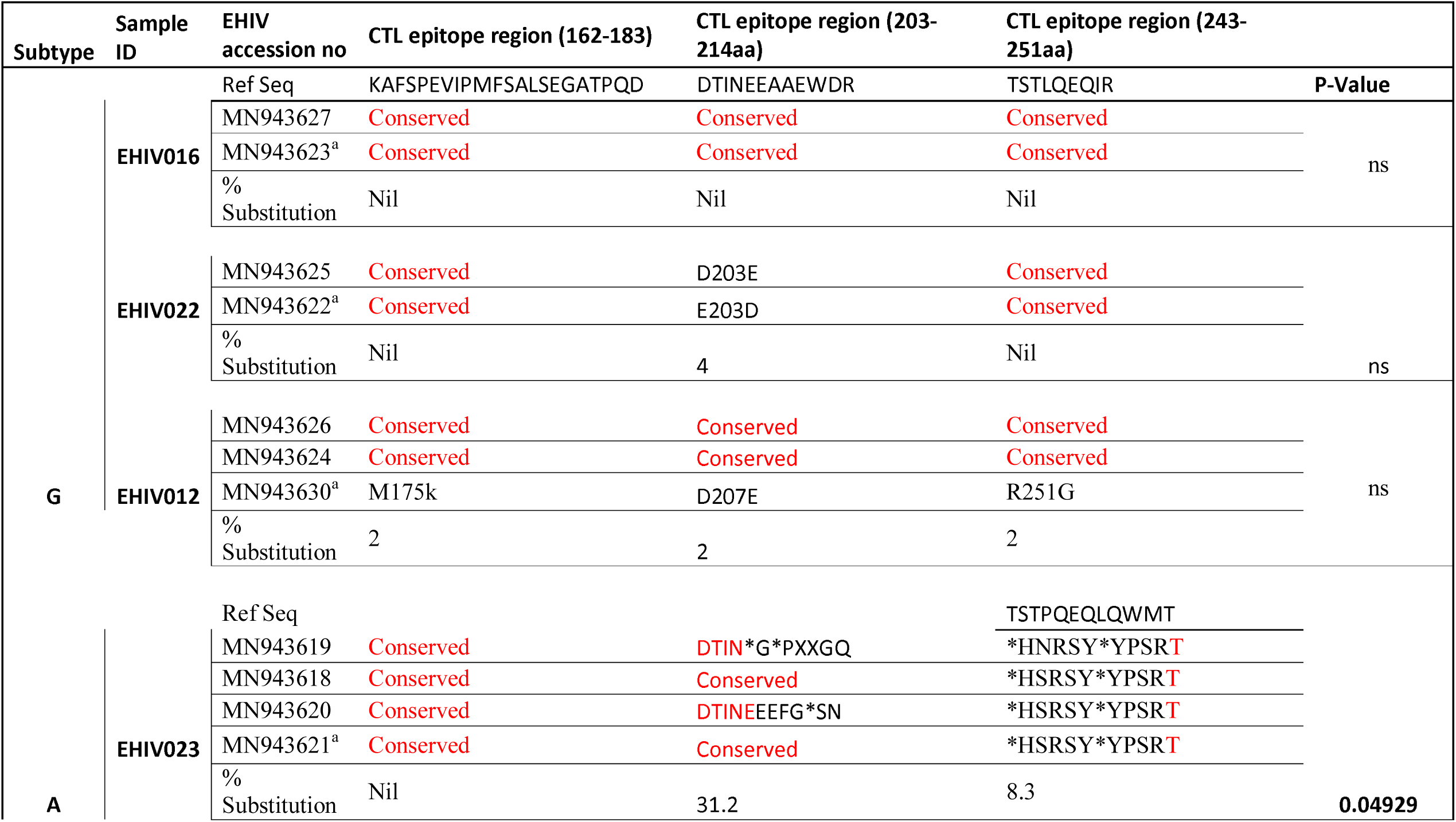
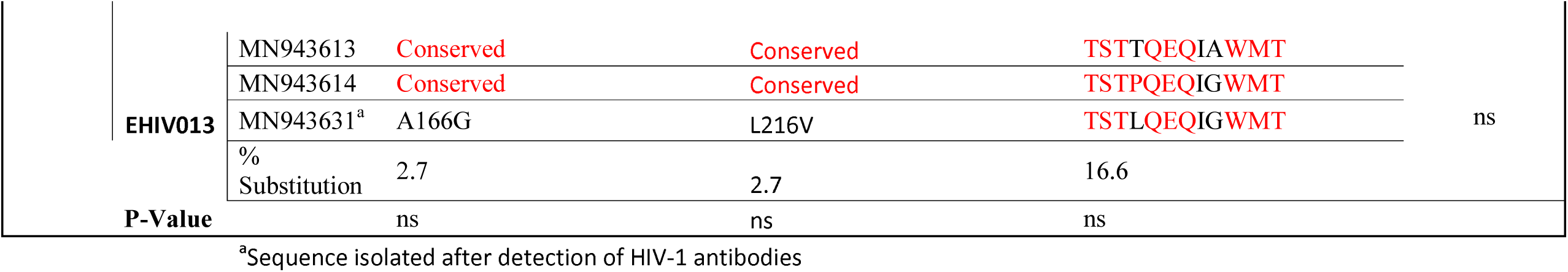
Substitutions associated with highly conserved sites in HIV-1 GAG gene isolated from 5 individuals followed from early HIV infection till detection of antibodies. Amino acid lengths of CTL epitope regions are indicated in parenthesis. Reference HIV-1 GAG sequences for subtypes G, A and CRF02-AG were downloaded from Los Alamos national HIV sequence database (www.hiv.lanl.gov/content/sequence/NEWALIGN/align.html) and were used for comparisons with similar subtypes obtained from persons at the early stages of HIV infection. ^a^Indicated accession number for sequences obtained from samples after detectable HIV-1 antibodies. Sequences that are not different from reference sequences (presence of the consensus amino acid) are indicated with the word Conserved. If an amino acid position is variable to the reference sequence, the letter and the position is indicated in the normal convention, e.g MN943625 sequence is variable at position 203 and it is indicated as D203E which means at position 203, D is substituted with E. When more than two amino acid positions are variable, the entire sequence is written against the accession number of the subtype. ^a^Reference sequence for subtype A CTL epitope region 243-251 is different from those of other subtypes and it is indicated. %Substitution was calculated by deducing the percentage of the ratio of variable positions against total amino acid positions for each sequence. Statistical tests of significance within and across CTL epitope regions for the subtypes studied were calculated using Kruskal-Wallis test, with P set at 0.05. ns=not significant. Significant values are indicated in bold values.

The mutation identified after detection of antibodies –E203D-in EHIV022 have been previously associated with escape[41]. EHIV012 had single aa substitutions each after detection of antibodies in the three CTL epitope regions studied. These substitutions were not previously observed. For HIV-1 subtype A samples, EHIV023 had very high substitution rates in CTL epitope region of 203-214aa (31.2%), although there were reversions by the time antibodies were detected. Amino acid substitutions were also observed in CTL epitope region spanning 243-251aa (8.3%). Significant differences in substitution rates before and after detection of antibodies across two of the three CTL epitope regions was observed for EHIV023 sample (see Table 4). Substitutions were more associated with the HIV-1 GAG gene sequenced after detection of antibodies for EHIV013 across the three CTL epitope regions studied. The two amino acid substitutions, A166G and L216V, observed in EHIV013 occurred after antibody detection and were not previously identified.

### Amino acid signature patterns in variable sites of HIV-1 GAG genes

Assessment of signature patterns in this study were limited to non-synonymous substitutions in which there were 100% aa replacements. In Table 5, these substitutions were compared between reference sequence and sequences isolated from individuals at the early stages of infection while in Tables 6 and 7, the substitutions were compared within sequences isolated per sample spanning early HIV infection till detection of antibodies. As shown in Table 5, non-synonymous substitutions were mostly observed in subtype G. However, a substitution, E105K, observed in CRF02_AG had been previously identified as a variant that did not recognize the HXB2 epitope[48]. Subtype A had a frame shift mutation at aa85-88. There were no substitutions in sequences isolated from sample EHIV016 while EHIV 012 had the highest number of substitution within Subtype G sequences (see Table 6). EHIV022 and EHIV 012 had substitutions within sequences at aa positions of 106 and 110-113.

**Table 5:** Frequency of non-synonymous substitutions in variable sites of HIV-1 GAG gene amino acids. Reference amino acid HIV-1 GAG sequences for subtypes G, A and CRF02-AG were downloaded from Los Alamos national HIV sequence database (www.hiv.lanl.gov/content/sequence/NEWALIGN/align.html) and were used as background sequences to query similar subtypes obtained from persons at the early stages of HIV infection using VESPA program (http://www.hiv.lanl.gov/content/sequence/VESPA/vespa.html) for variable signature amino acid residues. ^a^Indicated amino acid replacements were 100% substituted in the EHIV samples compared to reference sequences. These substituted residues are more likely explored as possible transmission and pathogenesis signatures. Residue replacements were restricted to these signatures with 100% substitutions to reduce, if not eliminate false discovery rate-adjusted P-values (Benjamin-Hochberg procedure). *indel in amino acid positions 85-88 introduced a frame shift mutation that terminated the computation of signatures for subtype A.

|  |  |  |  |  |  |  |  |  |  |
| --- | --- | --- | --- | --- | --- | --- | --- | --- | --- |
| **G** | aa position in<br>HIV-1 GAG gene | 16 | 21 | 34 | 68 | 70 | 103 | 107 | %sub |
|  | aa in Ref seq | L | V | Y | S | X | G | D |  |
|  | aa in EHIV<br>samples | X | M | N | G | Y | C | V | 2.3 |
| **CRF02**<br>**AG** | aa position in<br>HIV-1 GAG gene | 92 | 97 | 105 | 116 | 176 |  |  |  |
|  | aa in Ref seq | L | T | E | Q | T |  |  |  |
|  | aa in EHIV<br>samples | Q | K | K | I | K |  |  | 1.6 |
| **A\*** | aa position In<br>HIV-1 GAG gene | 89 |  |  |  |  |  |  |  |
|  | aa in Ref seq |  |  |  |  |  |  |  |  |
|  | aa in EHIV<br>samples | X |  |  |  |  |  |  |  |

These aa replacements were 100% substituted in the EHIV samples compared to Ref seq. \*Indel in aa positions 85-88 introduced a frame shift

**Table 6:**
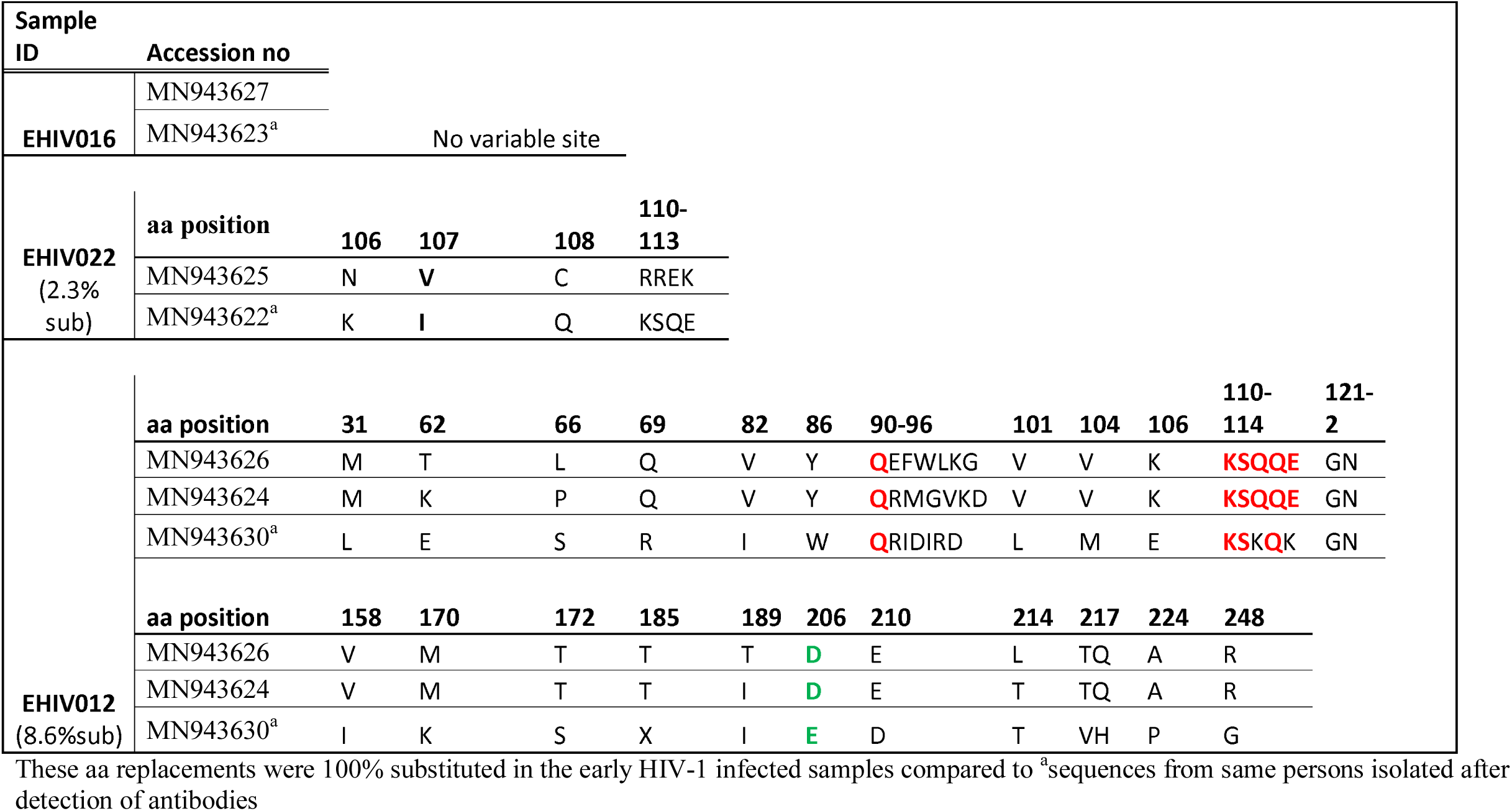
Frequency of non-synonymous substitutions in variable sites of HIV-1 Subtype G GAG gene isolated from 3 individuals followed up from early infection till detection of antibodies. Amino acid sequences obtained during early stages of HIV-1 infection were used as background sequences to query ^a^sequences obtained from same persons after antibodies detection using VESPA program (http://www.hiv.lanl.gov/content/sequence/VESPA/vespa.html) for variable signature amino acid residues. Indicated amino acid replacements were 100% substituted in the early HIV infected samples compared to sequences obtained after antibodies detection. These substituted residues are more likely explored as possible transmission and pathogenesis signatures. Residue replacement was restricted to these signatures with 100% substitutions to reduce, if not eliminate false discovery rate-adjusted P-values (Benjamin-Hochberg procedure).

**Table 7:** Frequency of non-synonymous substitutions in variable sites of HIV-1 Subtype A GAG gene isolated from 2 individuals followed up from early infection till detection of antibodies. Amino acid sequences obtained during early stages of HIV-1 infection were used as background sequences to query ^a^sequences obtained from same persons after antibodies detection using VESPA program (http://www.hiv.lanl.gov/content/sequence/VESPA/vespa.html) for variable signature amino acid residues. Indicated amino acid replacements were 100% substituted in the early HIV infected samples compared to sequences obtained after antibodies detection. These substituted residues are more likely explored as possible transmission and pathogenesis signatures. Residue replacement was restricted to these signatures with 100% substitutions to reduce, if not eliminate false discovery rate-adjusted P-values (Benjamin-Hochberg procedure).

| Sample ID | Accession no |  |  |  |  |  |  |  |  |  |  |  |  |  |
| --- | --- | --- | --- | --- | --- | --- | --- | --- | --- | --- | --- | --- | --- | --- |
| **EHIV023**<br>(4% sub) | **aa position** | **205** | **215-222** | **228-230** |  |  |  |  |  |  |  |  |  |  |
|  | MN943619 | I | LHPVHAGX | DKR |  |  |  |  |  |  |  |  |  |  |
|  | MN943618 | I | QG*LYPRC | PAX |  |  |  |  |  |  |  |  |  |  |
|  | MN943620 | T | NG*LYPRC | DKX |  |  |  |  |  |  |  |  |  |  |
|  | MN943621<sup>a</sup> | I | LHPVHAGX | DKR |  |  |  |  |  |  |  |  |  |  |
| **EHIV013**<br>(13.6%sub) | **aa position** | **15** | **28** | **30** | **41** | **49** | **60** | **62-62** | **66** | **69** | **72** | **76** | **82** | **86** |
|  | MN943613 | K | T | K | M | G | V | EQ | S | K | T | R | V | Y |
|  | MN943614 | S | K | R | L | S | I | DR | P | K | S | R | V | Y |
|  | MN943631<sup>a</sup> | A | K | R | L | G | L | EQ | S | R | S | K | I | W |
|  | **aa position** | **92-93** | **104** | **107** | **115** | **118** | **121** | **124** | **125-127** | **139** | **143** | **146-7** | **182** | **215** |
|  | MN943613 | EV | I | I | E | A | D | N | SKV | Q | V | AV | G | L |
|  | MN943614 | DV | I | M | P | A | D | S | SKV | Q | V | TL | Q | L |
|  | MN943631<sup>a</sup> | DI | M | I | V | T | A | S | *** | R | T | SM | Q | V |
|  | **aa position** | **218** | **223-224** | **228** | **243** | **248** | **252** | **260** | **263** | **267** |  |  |  |  |
|  | MN943613 | L | IA | I | T | A | G | D | R | I |  |  |  |  |
|  | MN943614 | V | VP | M | P | G | S | D | K | I |  |  |  |  |
|  | MN943631<sup>a</sup> | V | IP | M | L | G | S | E | K | V |  |  |  |  |

These aa replacements were 100% substituted in the EHIV samples compared to <sup>a</sup>Sequence isolated after detection of antibodies.

Lysine is the commonest aa used at position 106 (3/5) while a single substitution were observed for glutamate and asparagine. KSQ was the commonest aa usage in positions 110-113 (3/5), other aa in these positions were RRE (1/5) and KSK (1/5). In Table 7, non-synonymous substitutions associated with HIV-1 subtype A are described. EHIV013 had the highest number of substitutions spanning aa region 15 to 267. This is the largest number of substitutions found in this study. Samples EHIV023 and EHIV013 had substitutions within sequences at aa positions 215, 218 and 228. In position 215, Leucine was the highest aa used (4/7) while glutamine, asparagine and valine were present in one sequence each. Valine (4/7) and leucine (3/7) were the only aa used in position 218. Aspartate (3/7) was the highest aa used in position 228 while methionine (2/7), proline (1/7) and isoleucine (1/7) were also present in some sequences.

### Non synonymous substitutions associated with immune escape variants are more within epitopes outside the MHR

Although substitutions E105K (in CRF02AG), E203D (in subtype G) as well as K162R and A163G (in subtype A) are associated with immune escape and were observed as occurring within the MHR in this study, more substitutions previously associated with immune escape were found in regions outside the MHR. Out of the 5 individuals followed up, 2 had substitutions in GAG gene sites outside the MHR associated with immune escape strains (see Figures 2 and 3). Most mutations associated with previously described immune escape strains were identified after detection of antibodies in this study. Fourteen mutations in 16 HIV-1 GAG sites outside the MHR were identified before antibodies detection while 21 mutations in 23 HIV-1 GAG sites outside the MHR were identified after detection of antibodies. As shown in Figures 2 and 3, EHIV012 had three types of mutations before (L31M, L101V and S172T) and after (V82I, Y86W and F172S) detection of antibodies. H28K, M30R, A224P and A248G mutations were identified before and after detection of antibodies while V82I and Y86W mutations were identified only after detection of antibodies.

**Figure 2:**
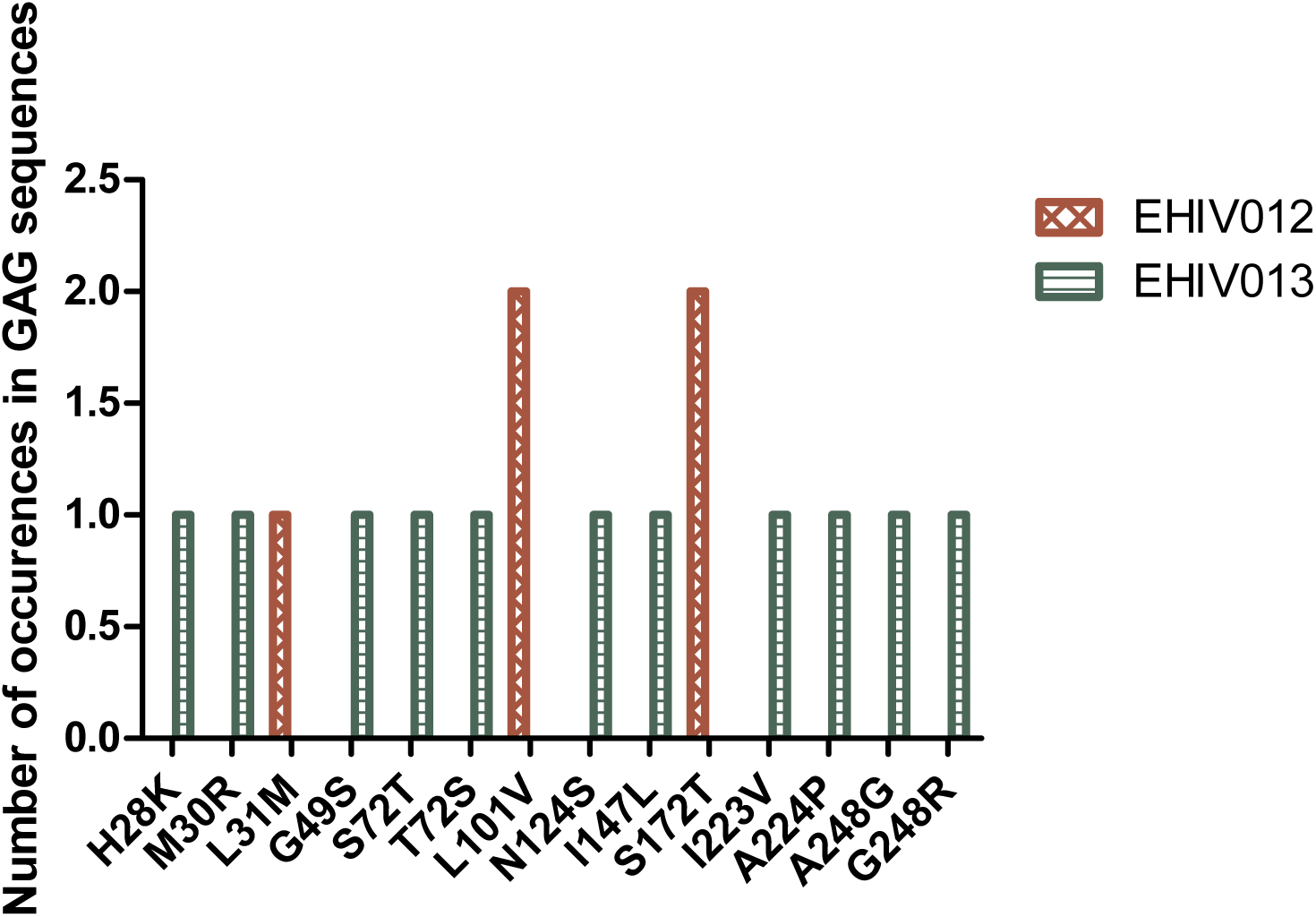
Distribution of mutations associated with escape in EHIV012 and EHIV013 before detection of antibodies. Three individuals had mutations previously associated with escape, diminished responses, non-susceptible forms etc. These mutations were compiled from the Los Alamos National Laboratory HIV Immunology Database for CTL/CD8+ Epitope Variants and Escape Mutations (https://www.hiv.lanl.gov/content/immunology/variants/ctl_variant.html). The list of all the identified mutations are presented in the Supplementary Table 1. Two individuals (EHIV012 and EHIV013) had mutations outside the GAG MHR while EHIV022 had a mutation corresponding to escape (Murakoshi) – E203D. Figure 3 shows the distribution of mutations associated with escape in EHIV 012(Red Bars) and EHIV013 (Green Bars). The number of occurrences of the mutations in GAG sequences are shown in the Y-axis while the X-axis shows aa mutations.

**Figure 3:**
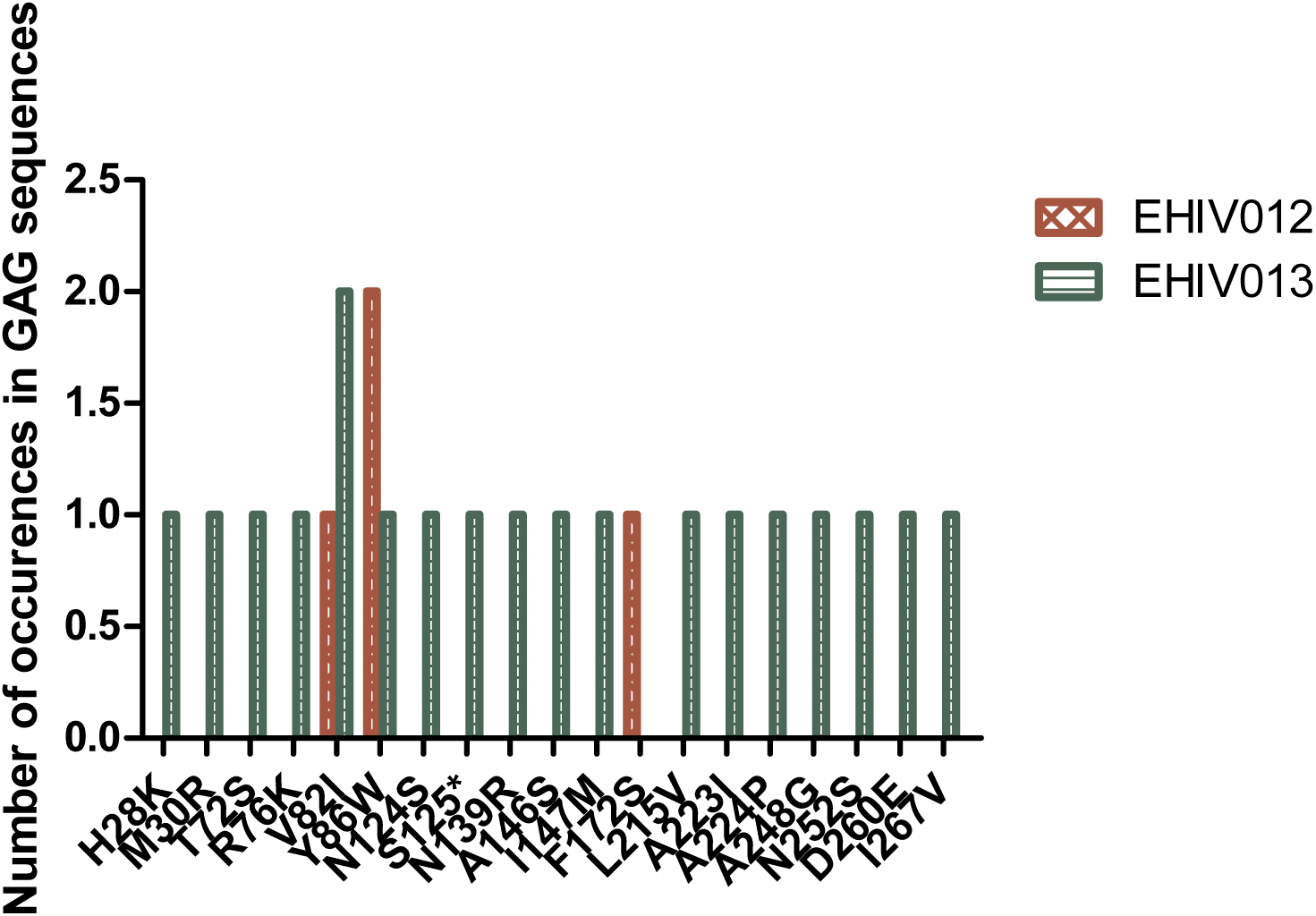
Distribution of mutations associated with escape in EHIV012 and EHIV013 after detection of antibodies. Three individuals had mutations previously associated with escape, diminished responses, non-susceptible forms etc. These mutations were compiled from the Los Alamos National Laboratory HIV Immunology Database for CTL/CD8+ Epitope Variants and Escape Mutations (https://www.hiv.lanl.gov/content/immunology/variants/ctl_variant.html). The list of all the identified mutations are presented in the Supplementary Table 1. Two individuals (EHIV012 and EHIV013) had mutations outside the GAG MHR while EHIV022 had a mutation corresponding to escape (Murakoshi) – E203D. Figure4 shows the distribution of mutations associated with escape in EHIV 012(Red Bars) and EHIV013 (Green Bars). The number of occurrences of the mutations in GAG sequences are shown in the Y-axis while the X-axis shows aa mutations.

### Serum creatinine concentration during longitudinal follow up

As shown in Figure 4, there was a significant difference (P<0.0044) in serum creatinine concentrations between early HIV-1 infection and detection of antibodies for the three individuals studied among the five persons followed up. EHIV 023 had lowest serum creatinine concentration at baseline (0.9mg/dl) and after antibody detection (0.8mg/dl) while EHIV 022 had the highest serum concentration of creatinine at baseline (1.1mg/dl) and after antibodies detection (1.0mg/dl). Sample EHIV016 had no changes in serum creatinine concentration from baseline until antibodies detection (1.0mg/dl).

**Figure 4:**
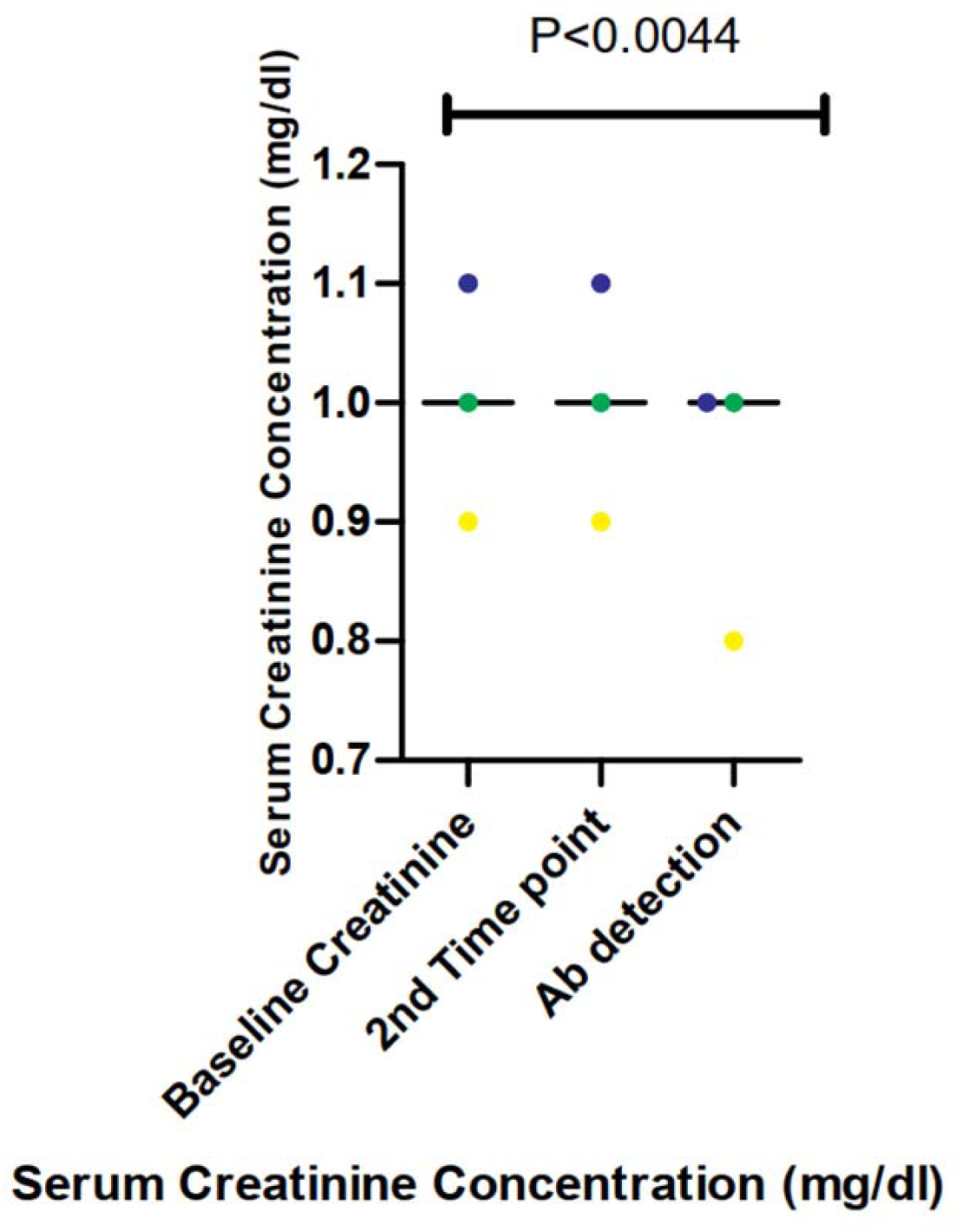
Serum creatinine concentration during longitudinal follow up of three individuals from early infection till detection of antibodies. The three individuals followed up are represented in blue, yellow and green. Serum creatinine levels for these individuals were measured in three time points, Baseline, second time point and after antibodies detection. Significant differences in the levels of serum creatinine (measured in mg/dl) observed across the three time points (P<0.0044) were calculated using 1-way ANOVA.

## Discussion

The results of this study show that diverse HIV-1 subtypes circulate in Nigeria, a West African country as subtypes A, G and CRF02_AG were identified. In our study, we identified 3 non synonymous substitutions within the MHR of HIV-1 GAG genes isolated from 10 early infected Nigerians. One substitution was however observed outside the MHR epitopes. Three (E105K, K162R and A163G) of these substitutions have been previously associated with immune escape[47,48]. These substitutions were associated with subtypes A and CRF02_AG. Most substitutions in subtype G within the MHR were associated with detection of antibodies while those of subtype A were associated with early HIV infection. Recent studies have showed that apart from immune escape mutations, other factors such as subtype differences and NEF mediated evasion of antibodies may account for replication and increased viremia during HIV-1 infection[49,50]. Fourteen mutations outside the MHR previously associated with immune escape were observed before detection of antibodies while 19 were observed after detection of antibodies in this study. This is a far cry from the four observed within the MHR. This finding is in consonance with previous reports that observed very limited non-synonymous substitutions in CTL epitopes within the MHR[7,28,51].

Gounder *et al*., 2015[52] however suggested that CTL escape immune variants observed during the early stages of HIV infection may be due to the transmitted virus as well as stochastic processes and not necessarily CD8 immune pressures while Roberts et al., 2015 reported that the average time before targeted epitope evolved an escape mutation was longer than two years[53]. We cannot fully corroborate this claim in our study because of our low sample size. However, since latent reservoir strains are established during the early stages of infection and are known to compose majorly of CTL immune escape strains[54,55], immune escape mutations may be incorporated more into the latent reservoirs irrespective of the mechanism with which they were generated. These latent reservoirs may require a broad CTL response for clearance as previously alluded[54]. Also, previous studies have showed that although escape mutants come with a fitness cost, this may be compensated for by HLA independent polymorphisms as well as other host-virus interaction factors[42]. Observations of CTL escape mutants even in the presence of antibodies in this study suggests that immune pressures by other immune cells other than CTL may aid the generation of CTL immune escape mutants. Mapping HIV immune epitopes in other regions of the genome will further clarify this hypothesis[27,56].

Universal HIV-1 vaccines are supposed to be broadly effective against all HIV-1 clades. However, immune escape strains encoding CTL epitopes outside the MHR may reduce the sensitivities of these vaccines. Also therapeutic HIV-1 vaccines are intended to be used after cART stoppage particularly against latent reservoir strains. However, as shown in this study, immune escape strains generated during the early stages of infection which are likely major constituents of the latent reservoir may lead to therapeutic vaccine failures[2,6,7,40]. Since immune escape occurs during the early stages of infection, it is imperative that cART commence early in order to reduce the reservoir size as well as the incorporation of immune escape variants[57,58]. Other studies have alluded to the fact that post treatment control may be possible if treatment begins early since blips observed after cART stoppage are largely due to immune escape variants incorporated into reservoir cells[59]. However, rapid and high magnitude CTL responses observed during the early stages of infection[11] may be affected by early treatment[58,60,61] which can in turn impair subsequent responses during cART stoppage in post treatment control trials. We have showed in this study that immune escape variants may be from those arising from CTL epitopes outside the MHR. However, functional and molecular studies on the nature and characteristics of HIV-1 strains in latent reservoirs need to be carried out to further ascertain our claims.

As observed previously in other studies, CTL epitope KAFSPEVIPMF was the most conserved[62,63]. This epitope has been associated with very low frequencies of CTL response selective pressures and has been a choice candidate for many T cell based HIV vaccines[2]. However, two substitutions previously associated with immune escape, K162R and A163G were observed in this epitope for Subtype A during the early stages of infection. On the other hand, CTL epitope DTINEEAAEWDR was associated with more substitutions, although most of these mutations reverted to wild type by the time antibodies were detectable. This phenomenon was observable in both subtypes A and G. It seems that although more mutations were observed during the early stages of HIV infection, reversions of these mutations occurred later on in infection. Previous studies have also associated reversions of CTL epitopes with the early stages of HIV infection[48]. A non-synonymous substitution, E203D, was observed in CTL epitope DTINEEAAEWDR. Selection of this epitope for immune escape strains have been previously described[41]. CTL epitope TSTLQEQIR was conserved for subtype G. However, this epitope was not prominent for subtype A sequences. Previous studies have documented that TSTLQEQIR may not be presented early and is associated with elite controllers[64,65]. Inclusion of this epitope in vaccines should induce strong CTL responses associated with viral load control and NEF gene down regulation. However, as observed in this study, the epitope is expressed by rare HLAs.

Several previously recognized immune escape substitutions were observed in this study. Majority of these substitutions emanated from epitopes outside the MHR. Although functional assays were not carried out to associate these epitopes with HLA polymorphisms, to the best of our knowledge, this is the first longitudinal study from West Africa on the natural history of HIV-1 especially as it relates to the kinetics of HIV-1 GAG CTL epitopes. We have also showed that numerous non-synonymous substitutions associated with CTL epitopes outside the MHR occur during the early stages of HIV-1 infection among HIV-1 subtypes and recombinant forms circulating in West Africa. It is important to state that these substitutions were identified from HIV-1 DNA sequences as against plasma RNA used in a previous similar study[52]. Proviral sequences have previously been associated with rare mutations on CTL epitopes[60,62]. These substitutions may have to be taken into consideration in future universal and therapeutic vaccines designs for HIV-1 strains circulating in West African countries[4,66]. More so that recent studies have showed that the major role of poorly recognized CTL epitopes is viral escape[67].

While some non-synonymous substitutions were observed before and after antibodies detection (H28K, M30R, A224P and A248G), some, such as V82I and Y86W were consistently identified after antibodies detection. V82I has been previously identified in studies among infected individuals[68,69]. This substitution among other things were associated with emergence of higher viral loads while Y86W was previously associated with HIV-1 clade B and E[70]. We as well as others have reported the high levels of creatinine among HIV infected Africans and African-Americans during the early stages of infection[43,44,71,72], findings from this present study show that these high levels may be linked to activation of HLA receptors on CD8 T cells during this stage of infection. Down regulation of this activation by the time antibodies appear and peak viremia subsides may correlate with reduction in creatinine levels.

In summary, we have showed that there is high genetic diversity of HIV-1 strains in Nigeria. Also, very high frequencies of non-synonymous substitutions occur in the HIV-1 GAG gene during the early stages of infection up until when antibodies become detectable. These substitutions include previous mapped CTL epitope immune escape mutants. We also observed different rates of HIV-1 GAG substitutions in the different subtypes especially during the early stages of infection. CTL immune pressure likely leave different footprints and signature patterns on HIV-1 GAG epitopes within and outside the MHR. This information is crucial for future vaccine designs for use in the West African region be it preventive or therapeutic.

## Data Availability

Data will be made available by the corresponding author upon request.

## Acknowledgements

BO, OD and GO conceptualized and designed the study. BO preformed experiments, analysed and interpreted the data as well as wrote the first draft of manuscript. OD and GO supervised the work and reviewed the manuscript.

